# COVID-19 infection and outcomes in a population-based cohort of 17,173 adults with intellectual disabilities compared with the general population

**DOI:** 10.1101/2021.02.08.21250525

**Authors:** Angela Henderson, Michael Fleming, Sally-Ann Cooper, Jill Pell, Craig Melville, Daniel MacKay, Chris Hatton, Deborah Kinnear

## Abstract

**Objectives:** To compare COVID-19 infection, severe infection, mortality, case-fatality, and excess deaths, among adults with intellectual disabilities and those without.

**Design:** Record-linkage of all adults recorded with intellectual disabilities in Scotland’s Census, 2011, and a 5% sample of other adults, to COVID-19 test results (Electronic Communication of Surveillance in Scotland), hospitalisations (Scottish Morbidity Record 01), and deaths (National Records of Scotland).

**Setting:** General population; 24^th^ January 2020 - 15^th^ August 2020

**Participants:** Successful linkage of 94.8% provided data on 17,173 adults with, and 195,859 without, intellectual disabilities.

**Outcomes:** Crude rates of COVID-19 infection, severe infection (hospitalisation/death), mortality, and case fatality; age-, sex- and deprivation-standardised severe infection and mortality ratios; annual all-cause mortality for 2020 and 2015-2019.

**Results:** Adults with intellectual disabilities had higher rates of COVID-19 infection (957/100,000 versus 513/100,000); severe infection (549/100,000 versus 237/100,000); mortality (259/100,000 versus 114/100,000); and case-fatality (30% versus 24%). Poorer COVID-19 outcomes remained after standardising for age, sex and deprivation: standardised severe infection ratio 2.59 (95% CI 1.80, 3.39) and mortality ratio 3.20 (95% CI 2.16, 4.25). These were higher among 55-64 year olds: 7.12 (95% CI 3.73, 10.50) and 16.16 (95% CI7.69, 24.63) respectively. Among adults with intellectual disabilities, all-cause mortality was only slightly higher in 2020 than the previous five years: standardised mortality ratios 2.49 (95% CI 2.17, 2.81) and 2.38 (95% CI 2.26, 2.49) respectively.

**Conclusions:** Adults with intellectual disabilities were more likely to be infected with COVID-19, and had worse outcomes once infected, particularly those under 65 years. Non-pharmaceutical interventions directed at formal and informal carers are essential to reduce transmission and all adults with intellectual disabilities should be immediately prioritised for vaccination regardless of age.

**Summary box:** *What is already known on this topic:*

- COVID-19 mortality is higher within multi-occupancy residences.
- Adults with intellectual disabilities may be at higher risk of COVID-19 mortality than other adults, but there are gaps in the evidence.
- COVID-19 case-fatality may be no different, or as much as 2.75 times higher in adults with intellectual disabilities compared with other adults.

*What this study adds:*

- Compared with general population adults, adults with intellectual disabilities were almost twice as likely to become infected with COVID-19, 2.3 times as likely to have severe infection, 2.3 times as likely to have COVID-19 mortality, and had 25% higher COVID-19 case-fatality.
- After standardising for age, sex and deprivation, people with intellectual disabilities were 3.2 times more at risk of covid-19 mortality and 2.6 times more at risk of severe infection relative to those with no intellectual disabilities
- Compared with general population adults, adults with intellectual disabilities had poorer outcomes among non-elderly age-groups particularly those aged 55-65 years, men, and those living in less-deprived neighbourhoods.
- Non-pharmaceutical initiatives are important for carers and care-provider organisations, and adults with intellectual disabilities should be prioritised in the national rollouts of COVID-19 vaccination programmes, regardless of age, sex, or neighbourhood deprivation.

## Introduction

The first case of COVID-19 was confirmed in the UK on 24^th^ January 2020 and a pandemic declared by the World Health Organisation (WHO) on 12^th^ March 2020. There is global concern that adults with intellectual disabilities may be at higher risk of death from COVID-19, but there are gaps in the evidence.(1) The WHO defines intellectual disabilities as impairments in adaptive, social, and intellectual functioning (IQ<70), requiring daily support, with the onset in the developmental phase (<18 years).(2) People with intellectual disabilities account for <1% of the global population,(3, 4) and about 0.5% of adults, (4) and they experience substantial health inequalities, including multimorbidity,(5) and premature and avoidable mortality,(6, 7) often from respiratory conditions.(8, 9) They are also more likely to live in congregate settings or be in receipt of social care (10); recent studies have reported high rates of COVID-19 mortality within multi-occupancy residences.(11) However, questions remain as to whether, compared with other people, people with intellectual disabilities are more likely to contract COVID-19, and whether they have more severe infections, and higher COVID-19 mortality.

Existing evidence is inconclusive and has limitations. Two studies have reported COVID-19 mortality and compared this with the general population, as far as data permitted, but did not report COVID-19 infection rates or COVID-19 case-fatality. In one non-peer reviewed study three data sources in England were used to identify adults with intellectual disabilities who definitely or possibly died from COVID-19 from 1^st^ February 2020 to 5^th^ June 2020.(12) Underestimates and uncertainty around figures were acknowledged in the report, due to limitations in data sources, and reporting. Their analysis of two of these data sources resulted in crude COVID-19 mortality of 240/100,000 population (2.3 times the general population) and 192/100,000 population (3.1 times the general population) respectively. From their analysis of data from registered care settings, all-cause deaths among adults with intellectual disabilities tripled during 10^th^ April 2020 - 15^th^ May 2020 compared with the 2 previous years. By comparison, the rates doubling over the same time period among the general population. (12) Welsh general practice records, 1^st^ March 2020 - 26^th^ May 2020, recorded 31 deaths from COVID-19 among people with intellectual disabilities,(13) which equated to a higher age-standardised COVID-19 mortality than in the general population.(13)

Four other studies reported COVID-19 case-fatality rates, though with biased samples, and conflicting results. Electronic medical records from 42 health care organisations (hospitals, primary care, and specialty treatment providers, typically large academic health centres and their affiliates; so possibly non representative), from 30 countries, up to 14^th^ May 2020, ascertained >30,000 patients with COVID-19 infections.(14) The authors found no difference in overall case-fatality rates between the 150 people with intellectual disabilities and those without, but reported possibly higher case-fatality rates among younger ages where deaths were generally less common.(14) A large USA study using private insurance claims, between 1^st^ April 2020 and 31^st^ August 2020, reported higher COVID-19 case-fatality among people with intellectual disabilities compared to those without (OR 2.75, 95% CI 1.66-4.56), especially at <70 years of age (OR 3.61, 95% CI 1.89-6.93).(15) However, these results may not be generalisable as they did not include people with public insurance or no insurance. A large self-selected sample of English general practices covering >4 million patients reported higher COVID-19 case-fatality among people with intellectual disabilities over weeks 2-20 of 2020 (OR 1.97, 95% CI 1.22-3.18).(11) A large study of people with intellectual and developmental disabilities living in about half of New York state’s residential settings were indirectly compared with the state’s general population up until 28^th^ May 2020. The study reported COVID-19 infection rate to be about 4 times higher, case-fatality almost double, and mortality rate 7.8 times higher.(16)

A prediction algorithm of risk of death following confirmed/suspected COVID-19 infection, was derived (24^th^ January 2020 - 30^th^ April 2020) and validated (1^st^ May 2020 - 30^th^ June 2020) using a large English primary care database of >8 million patients.(17) It reported higher fatality among adults with intellectual disabilities without Down syndrome (men: HR 1.36 (1.14-1.60), women: HR 1.36 (1.11-1.65)); and a further increase risk in the small sample of adults with Down syndrome (men: HR 9.80 (4.62-20.78), women: HR 32.55 (18.13-58.42)).(18) This led to the inclusion of Down syndrome, but not intellectual disabilities, to the clinically extremely vulnerable list used in the UK. (18)

It seems that adults with intellectual disabilities may be at greater risk of contracting COVID-19, and may be at greater risk of case-fatality, though evidence for both is currently limited. This study aimed to investigate in a whole-country adult population with intellectual disabilities, COVID-19 infection, severe infection, mortality, case-fatality, and excess deaths, compared with adults without intellectual disabilities, during the first wave of the COVID-19 pandemic (24^th^ January 2020-15^th^ August 2020).

## Methods

### Population, data sources, and record linkage

We used Scotland’s Census 2011 to identify all adults recorded as having intellectual disabilities and a random 5% sample of people not recorded as having intellectual disabilities. Census records were linked to COVID-19 laboratory tests, hospital admissions, and death registrations. Scotland’s Census, 2011, held by National Records of Scotland (NRS), provides detailed information on Scotland’s population, recorded on 27^th^ March 2011. NRS also collects data from death certificates, including date and International Classification of Diseases (ICD-10) cause of death. The Scottish Morbidity Record (SMR) 01 held by Public Health Scotland (PHS), records acute hospital admissions including dates of admission and discharge and ICD-10 diagnostic codes. Laboratory results from COVID-19 tests are stored electronically within PHS’s Electronic Communication of Surveillance in Scotland (ECOSS) database. These databases were linked together using a combination of deterministic and probabilistic matching. Personal identifiers from Census 2011 have previously been linked to the NRS Indexing Spine using probabilistic methods to create a read-through index for this dataset (19). This allows Census 2011 data to be linked to health data for research studies via the lookup held at NRS between the Community Health Index (CHI) number and the Indexing Spine. The CHI database is a population register of all Scottish residents in contact with NHS Scotland where each person has a unique ten digit CHI number.

The intellectual disabilities population was ascertained using Scotland’s Census 2011 which included the question ‘Do you have any of the following conditions which have lasted, or are expected to last at least 12 months? Tick all that apply.’ Respondents had 10 possible response options: (1) deafness or partial hearing loss; (2) blindness or partial sight loss; (3) learning disability (e.g. Down syndrome); (4) learning difficulty (e.g. dyslexia); (5) developmental disorder (e.g. autism spectrum disorder or Asperger syndrome); (6) physical disability; (7) mental health condition; (8) long-term illness, disease or condition; (9) other condition; (10) no condition. Those who selected option 3 were included in the intellectual disabilities cohort (as in Scotland, the term “learning disability” is synonymous with the international term “intellectual disabilities”).

We presented demographic characteristics for adults with and without intellectual disabilities; sex, age, and neighbourhood deprivation recorded at the time of the Census in 2011. Area neighbourhood deprivation status was derived from postcode of residence using the Scottish Index of Multiple Deprivation (SIMD) 2012 which is derived from 38 indicators across 7 domains (income, employment, health, housing, geographic access, crime and education, skills and training) using information collected for data zones of residence (median population 769). Adults were allocated to population deciles (1=most deprived; 10=least deprived). To reduce the risk of disclosing personally identifiable information on individuals, age was categorised as 18-54 years, 55-64 years, and ≥65 years, and SIMD was categorised into two groups: more deprived (deciles 1-5) and less deprived (deciles 6-10). People <18 years in 2011 were excluded from the analyses.

### Outcomes

The outcomes for this study were COVID-19 infection (positive COVID-19 test, hospitalisation for COVID-19, or death due to COVID-19); severe COVID-19 infection (hospitalisation for COVID-19 or death due to COVID-19); COVID-19 mortality; COVID-19 case-fatality (death from any cause among those who have had COVID-19 infection); and excess deaths (difference between average annual all-cause mortality rates 24/01-15/08 2015-2018 and all-cause death rate in 24/01/20 – 15/08/20).

Hospitalisation or death due to COVID-19 was defined as an ICD-10 code of U07.1 (confirmed COVID 19) or U07.2 (suspected COVID 19) in any primary or secondary diagnostic or cause of death position.

### Analyses

We had access to complete NRS death data up to 15th August 2020 and therefore investigated results over the period 24th January 2020-15th August 2020. Crude rates (per 100,000 people) were compared for those with and without intellectual disabilities using the number of people still alive within each group on 24th January 2020 as the respective denominators. Our crude outcomes included rates of COVID-19 infection, severe COVID-19 infection, COVID-19 mortality, and COVID-19 case-fatality. To take into account demographic differences between the groups with and without intellectual disabilities we then performed indirect standardisation using sex, age, and deprivation. We produced COVID-19 specific Standardised Mortality Ratios (SMRs) and COVID-19 specific Standardised hospitalisation/mortality Ratios for 2020. We then produced all-cause SMRs for deaths in 2020 and separately for deaths occurring over the previous 5 years. For each standardisation we used the 5% sample of the census population without intellectual disabilities as the standard population and compared the relevant age-sex-deprivation specific rates to the population with intellectual disabilities to ascertain expected and observed counts. We then repeated these analyses within sex, age, and deprivation subgroups standardising each time for the other two variables.

When calculating standardised ratios for deaths and admissions occurring in 2020 we used the respective denominator populations which included everyone in the original linked census cohort minus those who had died before Jan 24th 2020. When calculating standardised ratios for deaths between 2015 and 2019 we only counted deaths between 24th January and 15th August in each of the respective years in order to enable an accurate comparison with 2020. Subjects who had died before the 24th January in each of the respective years from 2015 through to 2019 were removed from the respective denominator populations contributing to the calculation. The 2015 denominator population for example included everyone in the original linked census cohort minus those who had died before Jan 24th 2015 and this was repeated for each subsequent year up to 2019. For all outcomes, age was calculated at time of event for those who had an event of interest (positive COVID-19 test, hospital admission, death) or age at 24th January in the respective year of interest for those people within the denominator population who did not have any events of interest.

### Patient and Public Involvement

There have been urgent calls from intellectual disabilities self-advocacy and third sector groups in Scotland to investigate the impact of COVID-19 on the population with intellectual disabilities, which this research addresses. There was no direct involvement of people with intellectual disabilities in this study. An accessible version of this report will be compiled to support equitable dissemination of these findings.

### Ethical approval

The programme of research, under which this research project sits, was approved by Scotland’s Public Benefit and Privacy Panel for Health (reference: 1819-0051), Scotland’s Statistics Public Benefit and Privacy Panel (1819-0051), and the University of Glasgow’s College of Medical, Veterinary, and Life Sciences Ethical Committee (reference: 200180081).

## Results

### Patient characteristics

Of the 293,869 people (24,264 with and 269,605 without intellectual disabilities) included in our Census 2011 cohort, 278,713 (94.8%) were successfully linked to the NRS Population Spine. The linkage rate was 92.9% (n=22,538) among people recorded as having intellectual disabilities and 95.0% (n=256,175) of the original 5% comparison sample with no intellectual disabilities. People <18 years old were subsequently excluded from the study (Figure 1) leaving a final cohort of 213,032 adults (17,173 adults with intellectual disabilities and 195,859 adults with no intellectual disabilities).

**Figure 1.**
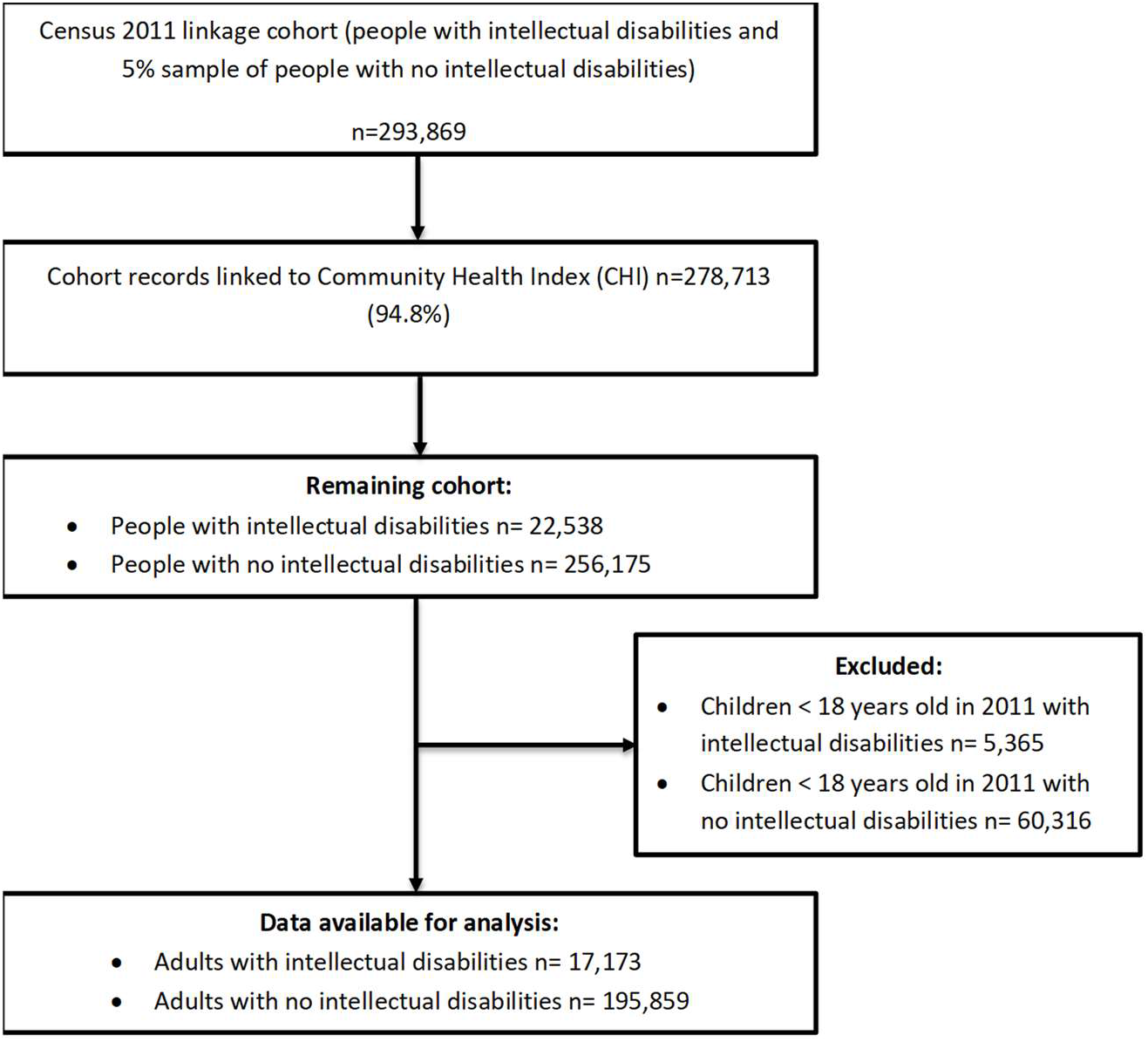
Participant flow diagram.

As expected, there were more men than women in the intellectual disabilities population, who were also younger, and more likely to live in deprived areas (Table 1).

**Table 1:**
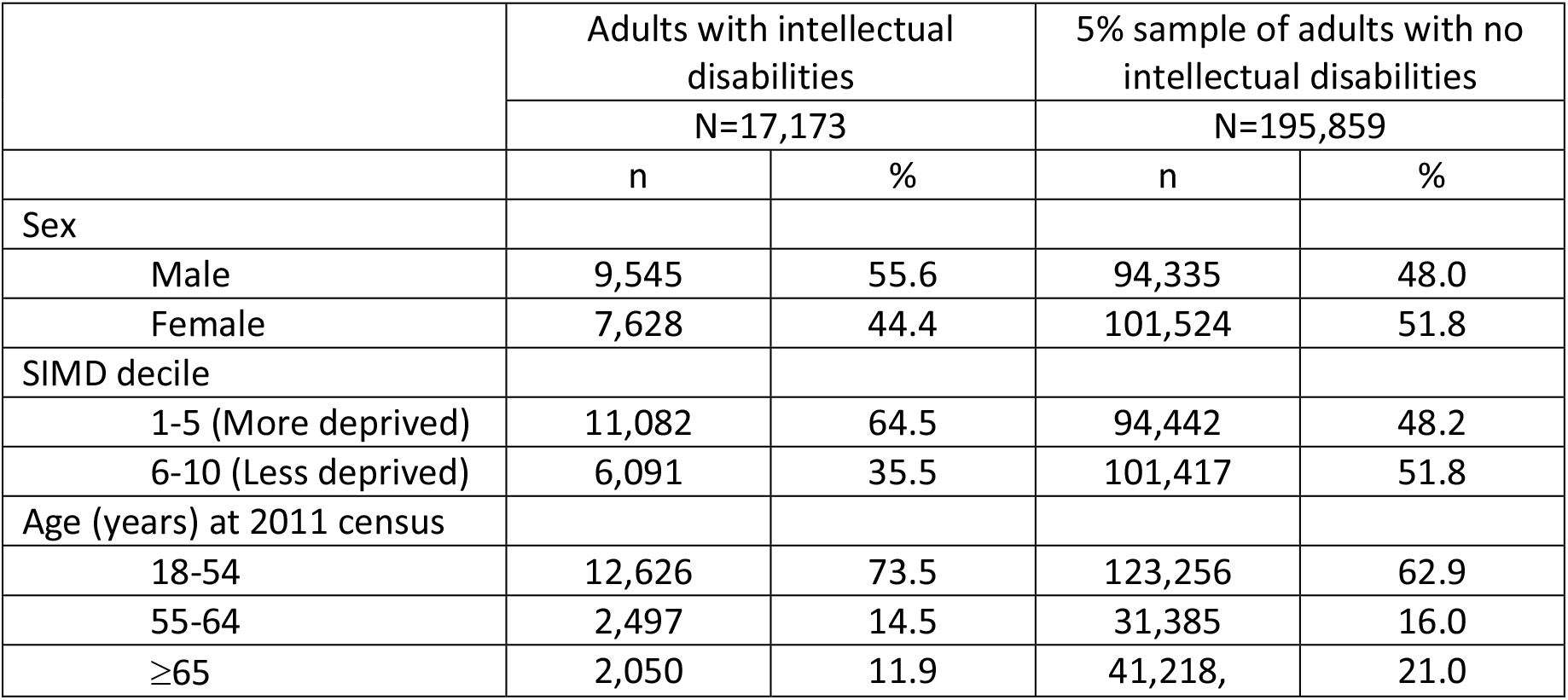
Characteristics of the study population

### Crude COVID-19 infection rates and outcomes

Compared to those without intellectual disabilities, adults with intellectual disabilities were almost twice as likely to become infected with COVID-19 (957/100,000 versus 513/100,000) and 2.3 times as likely to have severe infection resulting in hospitalisation or death (549/100,000 versus 237/100,000) or a fatal infection (259/100,000 versus 114/100,000) (Table 2). Following COVID-19 infection, people with intellectual disabilities were more likely to die 30% (95% CI 22%-38%) versus 24% (95% CI 21%-27%).

**Table 2:** Crude outcomes of study populations

|  | Adults with intellectual disabilities |  |  | 5% sample of adults with no intellectual disabilities |  |  |
| --- | --- | --- | --- | --- | --- | --- |
|  | N | Crude rate per 100,000 | 95% CI<sup>a</sup> | N | Crude rate per 100,000 | 95% CI<sup>a</sup> |
| COVID-19 |  |  |  |  |  |  |
| Infection<sup>b</sup> | 126 | 957 | 795, 1,119 | 892 | 513 | 480, 545 |
| severe infection<sup>c</sup> | 75 | 549 | 418, 661 | 412 | 237 | 214, 260 |
| Mortality<sup>d</sup> | 36 | 259 | 175, 344 | 199 | 114 | 99, 130 |

<sup>a</sup> 95% CIs calculated based on normal approximation
<sup>b</sup> positive COVID-19 test, hospitalisation for covid-19 or death from COVID-19
<sup>c</sup> hospitalisation for COVID-19 or death from COVID-19
<sup>d</sup> death from COVID-19 in the population

### Age-, sex-, deprivation-standardised COVID-19 outcomes

In 2020, the age-, sex-, deprivation-standardised ratio for severe COVID-19 infection was 2.59 (95% CI 1.80, 3.39) and for COVID-19 mortality was 3.20 (95% CI 2.16, 4.25), among adults with intellectual disabilities compared to those without (Table 3). The standardised ratios were slightly higher in men than women, and in less deprived areas. They were higher in people under 65 years of age and particularly high in the 55-64 year age group where the risk of severe infection, resulting in hospitalisation or death, was more than 7 times higher and the risk of death was over 16 times higher. (Table 3).

**Table 3.** Standardised* COVID-19 outcomes, overall and by sub-group

|  |  | Standardised hospitalisation/mortality ratio<sup>a</sup><br>(95% CI)<sup>c</sup> | Standardised mortality ratio<sup>b</sup><br>(95% CI)<sup>c</sup> |
| --- | --- | --- | --- |
| Overall |  | 2.59 (1.80, 3.39) | 3.20 (2.16, 4.25) |
| Sex | Male | 2.93 (1.84, 4.01) | 3.65 (2.19, 5.11) |
|  | Female | 2.08 (0.95, 3.21) | 2.57 (1.11, 4.02) |
| Deprivation | SIMD 1-5 (more deprived) | 2.27 (1.43, 3.11) | 3.06 (1.91, 4.22) |
|  | SIMD 6-10 (less deprived) | 3.75 (1.71, 5.79) | 3.71 (1.28, 6.13) |
| Age (years)<sup>d</sup> | 18-54 | 2.76 (1.05, 4.48) | 6.62 (2.03, 11.20) |
|  | 55-64 | 7.12 (3.73, 10.50) | 16.16 (7.69, 24.63) |
|  | ≥65 | 1.43 (0.68, 2.17) | 1.53 (0.73, 2.33) |

<sup>a</sup> Subgroup standardised hospitalisation/mortality ratios have been calculated based on fewer than 20 events in intellectual disabilities group and the results should therefore be treated with caution
<sup>b</sup> Subgroup standardised mortality ratios have been calculated based on fewer than 20 deaths in intellectual disabilities group and the results should therefore be treated with caution
<sup>c</sup> 95% CIs calculated based on normal approximation
<sup>d</sup> Age at time of event

### Excess overall mortality

Overall, the age-, sex-, deprivation-standardised all-cause mortality ratio for adults with intellectual disabilities was 2.38 (95% CI 2.26,2.49) over the five years prior to COVID-19, and only slightly higher at 2.49 (95% CI 2.17, 2.81) in 2020 (Table 4). In the sub-group analyses, the largest increase occurred in the 55-64 year age-group where the standardised all-cause mortality ratio increased from 4.26 (3.85, 4.66) between 2015 and 2019 to 5.09 (3.93, 6.25) in 2020. However, the confidence intervals still overlapped.

**Table 4.**
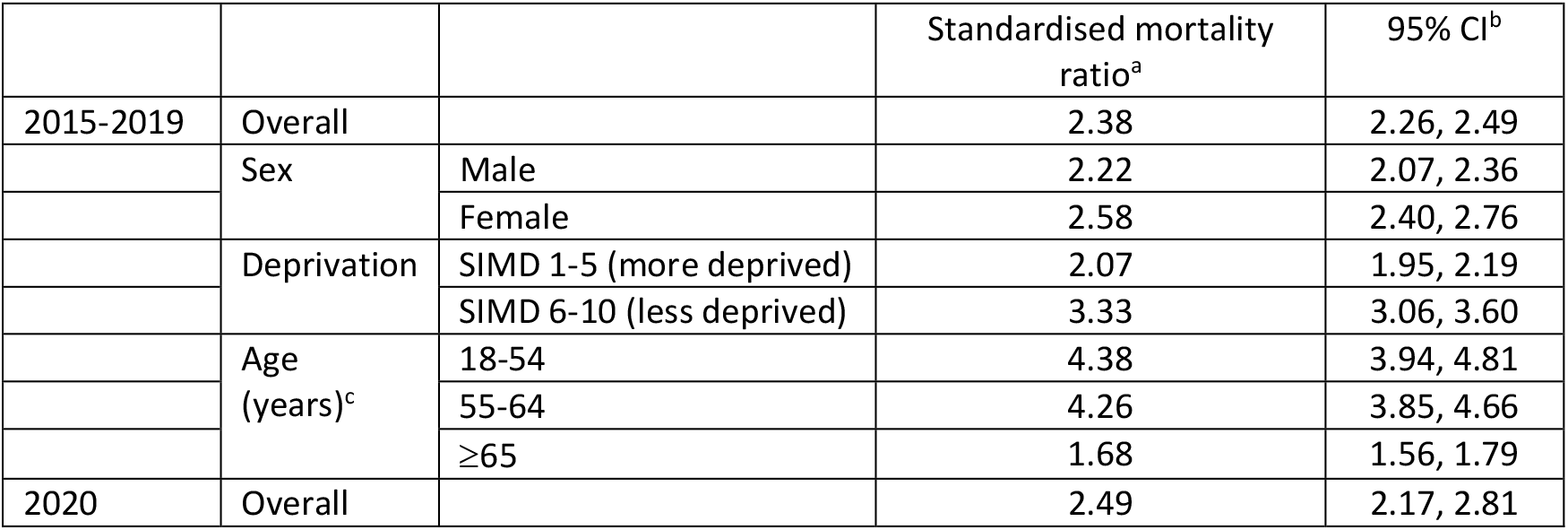
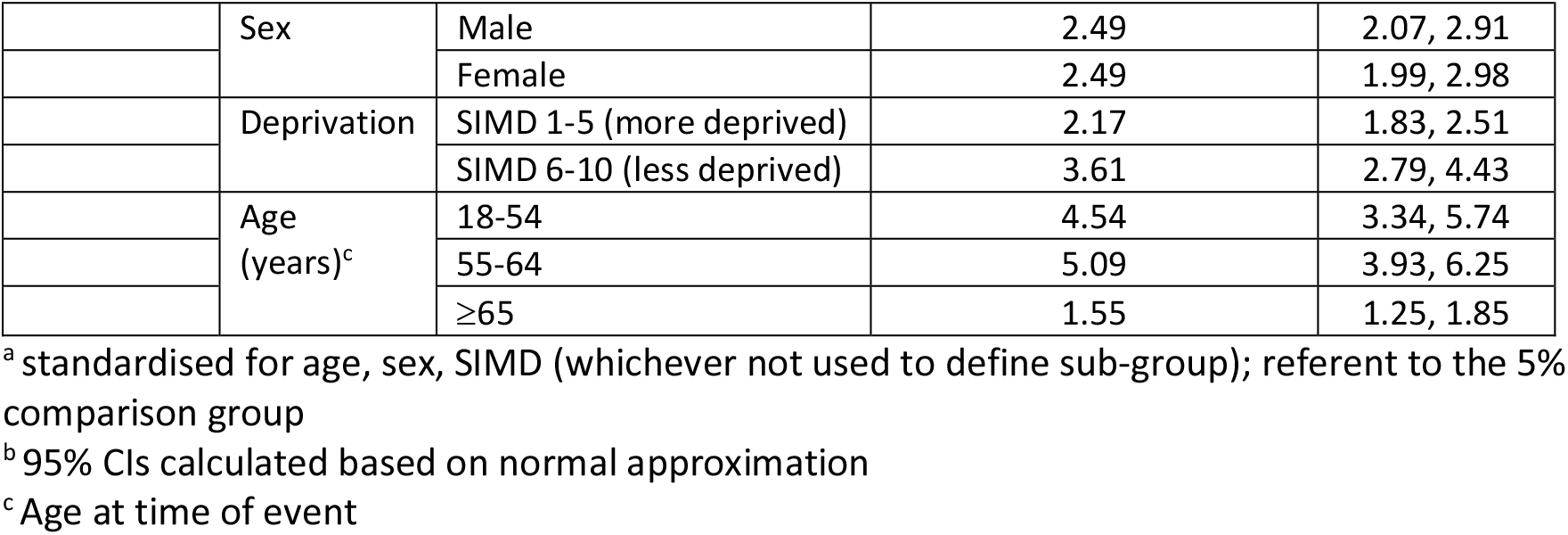
Age-, sex-, SIMD-standardised all-cause mortality, overall and by sub-group

|  |  |  | Standardised mortality ratio<sup>a</sup> | 95% CI<sup>b</sup> |
| --- | --- | --- | --- | --- |
| 2015-2019 | Overall |  | 2.38 | 2.26, 2.49 |
|  | Sex | Male | 2.22 | 2.07, 2.36 |
|  |  | Female | 2.58 | 2.40, 2.76 |
|  | Deprivation | SIMD 1-5 (more deprived) | 2.07 | 1.95, 2.19 |
|  |  | SIMD 6-10 (less deprived) | 3.33 | 3.06, 3.60 |
|  | Age (years)<sup>c</sup> | 18-54 | 4.38 | 3.94, 4.81 |
|  |  | 55-64 | 4.26 | 3.85, 4.66 |
|  |  | ≥65 | 1.68 | 1.56, 1.79 |
| 2020 | Overall |  | 2.49 | 2.17, 2.81 |
|  | Sex | Male | 2.49 | 2.07, 2.91 |
|  |  | Female | 2.49 | 1.99, 2.98 |
|  | Deprivation | SIMD 1-5 (more deprived) | 2.17 | 1.83, 2.51 |
|  |  | SIMD 6-10 (less deprived) | 3.61 | 2.79, 4.43 |
|  | Age (years)<sup>c</sup> | 18-54 | 4.54 | 3.34, 5.74 |
|  |  | 55-64 | 5.09 | 3.93, 6.25 |
|  |  | ≥65 | 1.55 | 1.25, 1.85 |

<sup>a</sup> standardised for age, sex, SIMD (whichever not used to define sub-group); referent to the 5% comparison group
<sup>b</sup> 95% CIs calculated based on normal approximation
<sup>c</sup> Age at time of event

## Discussion

### Principal findings and interpretation

This is the first comprehensive study to investigate COVID-19 infection, severe infection, mortality, case-fatality, and excess mortality among a whole country’s population of adults with intellectual disabilities compared with a representative sample of the general population. Overall, adults with intellectual disabilities were both more likely to be infected with COVID-19 and had worse prognosis once infected. They were twice as likely to become infected with COVID-19, 2.3 times as likely to have severe infection resulting in hospitalisation or death, and following COVID-19 infection, had a case-fatality of 30% compared with 24% in the general population. The risk of severe or fatal COVID-19 infection, relative to the general population, was higher among non-elderly age-groups and particularly high in those aged 55-64 years of age. This highlights the importance of action to reduce COVID-19 infection and mortality risks for adults with intellectual disabilities who are in younger as well as older age groups. The higher risk, relative to the general population, among those living in less deprived areas may simply reflect the lower absolute risk among the general population in these areas. The overall risk of dying from any cause was already higher among adults with intellectual disabilities prior to the COVID-19 pandemic. The net effect of COVID-19 is a slight, non-significant increase in excess deaths from any cause.

In normal daily activities, people with intellectual disabilities come into contact with numerous others including: residents in shared accommodation, shared care-givers, friends and families, and contacts in work and day placements. People with intellectual disabilities have higher rates of multi-morbidity resulting in more frequent contact with health-care workers. Therefore, even during “lock-down,” their contacts are likely to remain higher than the rest of the population. This highlights the importance of non-pharmaceutical interventions, such as social distancing, face coverings, and hand hygiene in minimising their risk of infection.

### Comparison with previous studies

Previous studies have suggested variable COVID-19 outcomes for people with intellectual disabilities. These report COVID-19 mortality rates 2-3 fold higher than the general population,(12, 13) no difference in case-fatality,(14) overall case-fatality with OR=2.75 or OR=3.61 in the <70 year olds,(15) and all-cause mortality in those with known COVID-19 with OR=1.97.(11) A more comprehensive study on adults with intellectual disabilities, including only those in residential settings (compared with whole-community general population) reported COVID-19 infection rates to be about 4 times higher, case-fatality to be almost double, and mortality rates to be 7.8 times higher.(16) In our study of a whole nation’s adult population with intellectual disabilities, we found they were twice as likely to become infected with COVID-19, with a case-fatality 25% higher than in the general population, and COVID-19 mortality was 2.3 times higher than in the general population.

Poorer COVID-19 outcomes in younger adult age groups compared with the general population were expected and have been previously suggested.(12, 14) This is in view of premature deaths resulting in people with milder intellectual disabilities and fewer co-morbidities surviving into old age, and therefore more closely resembling the elderly general population. This difference in the age profile of COVID-19 mortality is important given the current prioritisation of vaccinations in the UK on those in older age groups, which will potentially lead to increased levels of potentially preventable COVID-19 mortality in younger people with intellectual disabilities who are not vaccinated.

To our knowledge, only one study to date has investigated adults with intellectual disabilities with and without Down syndrome, finding that Down syndrome predicted time to death from COVID-19 to a greater extent than intellectual disabilities without Down syndrome, but was based on few people with Down syndrome in the dataset, and has not been replicated.(17) People with Down syndrome form less than a fifth of all adults with intellectual disabilities. The study also found that intellectual disabilities without Down syndrome predicted time to death.(18) Given the evidence presented from our study, along with the pre-existing literature, serious consideration now should be given to the addition of intellectual disabilities, as well as Down syndrome, to the clinically extremely vulnerable list.(18)

### Strengths and limitations

The study was large and included the entire country’s adult population with intellectual disabilities, as well as a representative proportion of adults in the general population, thereby reducing bias. Presence of intellectual disabilities was sought on everyone. There was a high, 94%, response rate. Importantly, intellectual disabilities was differentiated from specific learning disabilities (such as dyslexia), and autism. The question on intellectual disabilities was subject to cognitive question testing prior to use, to ensure that it accurately captured the condition, and was acceptable to the population. Record linkage was successful on 94.8% and provided data on a wide range of outcomes.

COVID-19 testing data, extracted from the ECOSS system for the period investigated (24th January 2020-15th August 2020), may be an underestimate of true community incidence of COVID-19 infection rates due to limited COVID-19 testing during the first wave of the pandemic, although there is no reason to suspect differences in testing rates between the adults with, and without, intellectual disabilities. Case fatality rates were high in both groups and are likely to be an overestimate due to lack of testing of less severe (asymptomatic/mild symptoms) infections, particularly in the early stages of the pandemic. We did not have information on ability level, Down syndrome, or living circumstances, data on other risk factors for COVID-19 such as comorbidities which are more common in adults with intellectual disabilities than other people, or information on health-care access which previously has been documented as being poorer for this population. In the sub-group calculations of standardised ratios of severe and fatal COVID-19 infections some of the cells contained less than twenty events. The Office for National Statistics (ONS) advises that, in such situations, the derived rates should be interpreted as having low reliability.(20) We used routinely collected data, and cause of death data is infrequently verified by post-mortem.

### Implications for clinicians and policymakers

Non pharmaceutical interventions, such as access to personal protective equipment, are even more important for carers and care-provider organisations supporting people with intellectual disabilities, in minimising the transmission of Covid-19, than the rest of the population. Our findings are important also for policy-makers, clinicians, and public health physicians to make evidence-based decisions about targeting preventive measures such as shielding, surveillance strategies, criteria for testing, and prioritisation for vaccination. These are relevant for all adults with intellectual disabilities, regardless of age, sex, or extent of deprivation of the neighbourhood they reside within. The age cut-offs used in the general population for prioritising COVID-19 vaccination should not be applied to adults with intellectual disabilities who are a higher risk even at younger ages.

## Data Availability

No data are available.

## Acknowledgments

The authors would like to acknowledge the support of the eDRIS Team (Public Health Scotland) and National Records of Scotland for their involvement in obtaining approvals, provisioning and linking data, and the use of the secure analytical platform within the National Safe Haven. This work uses data provided by patients and collected by the NHS as part of their care and support.

